# Enhancing COVID-19 Case Forecasting in the United States: A Comparative Analysis of ARIMA, SARIMA, and RNN Models with Grid Search Optimization

**DOI:** 10.1101/2024.03.04.24303713

**Authors:** Samira Nichols, Saina Abolmaali

## Abstract

The COVID-19 pandemic has resulted in a substantial number of fatalities in the United States since its onset in January 2020. In an effort to mitigate the spread of this highly infectious disease, a range of measures, including social distancing, mask-wearing, lockdowns, and vaccination campaigns, have been implemented. However, despite these extensive efforts, the persistent transmission of the virus can be attributed to a combination of vaccine hesitancy among certain individuals and the emergence of new viral strains. To effectively manage the ongoing pandemic, healthcare providers and government officials rely on infectious disease modeling to anticipate and secure the necessary resources. Accurate short-term case number forecasting is of paramount importance for healthcare systems.

Since the beginning of the pandemic, numerous models have been employed to forecast the number of confirmed cases. In this study, we undertake a comparative analysis of six time-series techniques: Simple Moving Average (SMA), Exponentially Weighted Moving Average (EWMA), Holt-Winters Double Exponential Smoothing Additive (HWDESA), Autoregressive Integrated Moving Average (ARIMA), Seasonal Autoregressive Integrated Moving Average (SARIMA), and Recurrent Neural Network (RNN), with regard to their modeling and forecasting capabilities. SMA, EWMA, and HWDESA were employed for predictive modeling, while the ARIMA, SARIMA, and RNN models were utilized for case number forecasting. A comprehensive grid search was carried out to determine the optimal parameter combinations for both the ARIMA and SARIMA models. Our research findings demonstrate that the Holt-Winters Double Exponential model outperforms both the Exponentially Weighted Moving Average and Simple Moving Average in predicting the number of cases. On the other hand, the RNN model surpasses conventional time-series models such as ARIMA and SARIMA in terms of its forecasting accuracy. The finding of this study emphasizes the importance of accurately predicting the number of COVID-19 cases, given the substantial loss of lives caused by both the virus itself and the societal responses to it. Equipping healthcare managers with precise tools like Recurrent Neural Networks (RNNs) can enable them to forecast future cases more accurately and enhance their preparedness for effective response.

## 1 Background

As of June 2, 2023, the United States has reported a total of 103,436,829 confirmed cases of COVID-19, according to data from the Centers for Disease Control and Prevention (CDC) [1]. The unpredictable nature of the virus has necessitated the continued use of quarantine as a primary response to curb its spread. Throughout the pandemic, various strategies have been implemented to control the disease’s transmission. One crucial strategy involves the utilization of statistical and mathematical modeling to monitor outbreaks and make informed decisions.

Time-series analysis stands out as an invaluable tool for modeling and forecasting the spread of COVID-19. These techniques have been widely applied to analyze time-dependent medical data, enabling predictions about the pandemic’s trajectory. For instance, Abolmaali and Shirzaei [2] employed linear regression, logistic regression, and the Susceptible, Infected, and Recovered (SIR) model to forecast the number of COVID-19 cases in four different U.S. states.

In addition to these traditional models, machine learning and artificial intelligence methods have been harnessed for predicting the spread of COVID-19. These models are trained on large historical datasets to forecast future case numbers and transmission rates, guiding the decisions of healthcare providers and policymakers. Furthermore, they assist in evaluating the effectiveness of intervention strategies like lockdowns and vaccination campaigns.

Lai [3] utilized Box-Jenkins and Autoregressive Integrated Moving Average (ARIMA) models to analyze SARS infection rates, while Martinez et al. [4] applied the Seasonal Autoregressive Integrated Moving Average (SARIMA) model to study dengue incidence, highlighting the effectiveness of these models in modeling infectious diseases.

In other studies, Roy et al. [5] used an ARIMA model to predict COVID-19 spread in India, and Tandon et al. [6] employed an ARIMA model to forecast short-term case increases in India. Koyuncu et al. [7] used the SARIMA model to forecast the impact of COVID-19 on maritime container throughput. Ceylan [8] employed ARIMA and SARIMA models to estimate COVID-19 prevalence in Italy, France, and Spain.

ARIMA models have been a common choice in many studies for predicting infectious disease patterns, including COVID-19. For instance, Bayyurt et al. [9], Yang et al. [10], Benvenuto et al. [11], Perone et al. [12], Ilie et al. [13], Singh et al. [14], and Sahai et al. [15] have effectively used ARIMA models to forecast COVID-19 transmission in different countries. These studies have demonstrated that ARIMA models are potent tools for forecasting infectious disease spread and can provide valuable insights for healthcare providers and policymakers. Several studies have explored death rates across various age groups. Notably, Jacobson et al. [16] found a statistically significant decrease in the risk of non-COVID-19-related deaths among the youngest age cohorts. Ludden et al. [17] utilized preliminary mortality data from the CDC to calculate mortality risks for 22 subgroups categorized by age and sex in 2021, and then compared them to the period from 2015 to 2019 using odds ratios. They concluded that among individuals aged 25 to 54, Year 2 (April 2021 to March 2022) exhibited a higher mortality rate compared to Year 1 (April 2020 to March 2021), whereas the overall mortality risks for individuals aged 65 and above decreased. In addition to the conventional time series forecasting methods mentioned earlier, Recurrent Neural Networks (RNNs) have proven to be powerful forecasting tools. This is underscored by their remarkable success in the winning strategy of the recent M4 competition [18, 19].

In this paper, our objective is to explore the application of time-series models for modeling and forecasting the prevalence of COVID-19 cases in four U.S. states: Alabama, Massachusetts, California, and Washington. We will specifically investigate six different time-series models: Simple Moving Average (SMA), Exponentially Weighted Moving Average (EWMA), Holt-Winters Double Exponential Smoothing Additive (HWDESA), and Recurrent Neural Network (RNN) to model the prevalence of confirmed and deceased COVID-19 cases. In the subsequent phase, we will employ Autoregressive Integrated Moving Average (ARIMA), Seasonal Autoregressive Integrated Moving Average (SARIMA), and Recurrent Neural Network (RNN) to forecast the number of infected individuals.

This comprehensive analysis across multiple states will yield a nuanced understanding of COVID-19’s spread in diverse U.S. regions. Furthermore, by comparing the performance of various time-series models, we can identify the most effective ones for modeling and forecasting the pandemic’s dynamics. This holistic approach will offer a more comprehensive perspective on the pandemic and provide valuable insights for policymakers.

## 2 Method

### 2.1 Data

The Johns Hopkins COVID-19 data repository is widely acknowledged as the most reliable source of information for COVID-19 statistics [20]. This repository provides comprehensive time-series data, encompassing daily counts of confirmed cases, fatalities, and recoveries.

Given the extensive and intricate nature of COVID-19 data, and recognizing that a thorough analysis of all U.S. states is beyond the scope of this paper, we have narrowed our focus to four specific states: Alabama, Washington, California, and Massachusetts. We’ve chosen to examine data from March 2020 to October 2023. These states were deliberately selected from various geographical regions, ensuring a comprehensive representation of the pandemic’s progression across diverse areas. This narrowed scope allows for a detailed examination of COVID-19’s dynamics in different parts of the country, facilitating a robust comparison of the various time-series models employed in this study.

### 2.2 Time series models

In this section, we will provide a brief overview of the time-series models that we will employ for modeling and forecasting the spread of COVID-19.

#### Simple Moving Average (SMA)

The Simple Moving Average (SMA) is a frequently employed technique for modeling time-series data. It involves computing the average of the data over a fixed number of periods, treating all data points with equal weight. The SMA is calculated using the equation presented in Eq. 1, where *C*_*t*_ represents the cumulative count of infected and deceased cases in period *t* and *n* denotes the length of the moving average time span (e.g., the number of months).

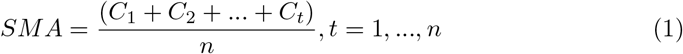

The Simple Moving Average (SMA) is a straightforward and easily comprehensible method for modeling time-series data, which is why it finds extensive application across various domains, including finance, economics, and epidemiology. In our study, we will employ the SMA technique to model the cumulative counts of both infected and deceased cases of COVID-19 in the four aforementioned states.

It’s important to acknowledge that while SMA is a valuable tool, it has limitations. SMA assigns uniform weight to all data points, making it less suitable for scenarios where the data exhibits non-stationarity, trends, seasonality, or other intricate patterns. Nevertheless, SMA serves as an excellent initial step in grasping the data’s overall trend and provides a benchmark against which we can evaluate the performance of more sophisticated models.

#### Exponentially Weighted Moving Average (EWMA)

The Exponentially Weighted Moving Average (EWMA) is a modification of the Simple Moving Average (SMA) method, distinguished primarily by its approach to incorporating data points into the moving average calculation. In contrast to SMA, which gives equal weight to all data points, EWMA employs weighted assignments. In our study, we assign a higher weight to the most recent observations. This means that the most recent data points exert a more pronounced influence on the moving average in comparison to historical data points. The calculation of EWMA is determined by the equation presented in Eq. 2. In this equation, *C*_*t*_ represents the cumulative count of infected and deceased cases in period *t* and *α* is a smoothing parameter within the range of 0 to 1. The parameter *α* plays a pivotal role in determining the weighting of data points; a higher *α* value accentuates the influence of recent data points, while a lower *α* value places greater emphasis on historical data points.

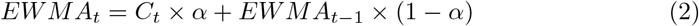

EWMA is a highly effective approach, particularly in situations where the data exhibits non-stationarity, which implies the presence of trends or seasonality, and where the data dynamically evolves over time. Given that recent data points carry greater weight, EWMA excels at adapting the moving average model in real-time to mirror shifts in the data. Additionally, it aids in attenuating noise within the data.

In our study, we will apply the EWMA technique to model the cumulative counts of both infected and deceased cases of COVID-19 in the four states under investigation. Subsequently, we will evaluate its performance relative to the SMA method.

### Holt-Winters Double Exponential Smoothing Additive (HWDESA)

The Holt-Winters Double Exponential Smoothing Additive (HWDESA) method stands as a widely adopted approach for modeling time-series data characterized by the presence of both seasonality and trend components. This method comprises three core equations, each addressing different aspects of the time series.

The level equation (Eq. 3) computes the series’ average value, offering a weighted average between the seasonally adjusted observation and the non-seasonal forecast. The trend equation (Eq. 4) quantifies the upward or downward movement within the series. Lastly, the seasonality equation (Eq. 5) captures the recurring seasonal patterns within the series by incorporating a weighted average between the current seasonal index and the seasonal index from the previous cycle of the same season.

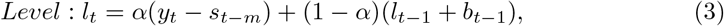

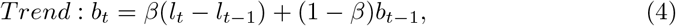

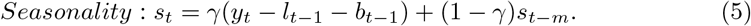

Within these equations, *α, β*, and *γ* represent smoothing parameters crucial for determining the weightings applied to the data. The selection of their optimal values hinges on minimizing errors, specifically by minimizing the sum of squared errors. This often involves employing techniques like grid search or other optimization methods to arrive at the best parameter values.

In our study, we will implement the Additive Holt-Winters method to model the cumulative counts of COVID-19 infections in the four states under investigation. Subsequently, we will assess its performance in comparison to the SMA and EWMA methods.

#### Autoregressive Integrated Moving Average (ARIMA)

The Autoregressive Integrated Moving Average (ARIMA) model belongs to a class of models designed to elucidate autocorrelation within time-series data. It is extensively employed for modeling and forecasting time series that can be rendered stationary through differencing. This model comprises three key parameters: Auto Regression (AR), Integrated (I), and Moving Average (MA), collectively defining an ARIMA model denoted as ARIMA(p, d, q). Here, p signifies the order of the Auto Regression, d represents the degree of differencing necessary to achieve stationarity in the time series data, and q denotes the order of the Moving Average component. The ARIMA(p, d, q) model is defined as follows:

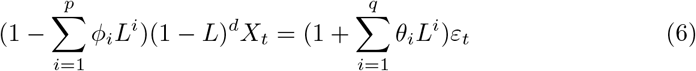

Where L is the lag operator, *ϕ*_*i*_ and *θ*_*i*_ are the parameters of the model, *X*_*t*_ is the time series, and *ε*_*t*_ is the error term.

The selection of appropriate values for the p, d, and q parameters is of paramount importance to ensure that the model accurately captures the underlying data patterns and provides precise forecasts. To determine these parameters, we employed a grid search approach and supplemented it with the Dickey-Fuller and the Canova-Hansen tests to determine the optimal differencing terms. The selection of the best combination of p, q, and d was guided by evaluating various ARIMA models using metrics such as the Akaike Information Criterion (AIC), Root Mean Square Error (RMSE), and Mean Square Error (MSE). These metrics served as performance indicators across different states.

In this study, ARIMA models were employed to model and forecast the counts of confirmed and deceased COVID-19 cases in the four states under investigation.

#### Seasonal Autoregressive Integrated Moving Average (SARIMA)

SARIMA, which stands for Seasonal Autoregressive Integrated Moving Average, is a category of models specifically tailored to account for seasonality within time series data. These models are essentially ARIMA models with an added seasonal component, rendering them capable of handling seasonal patterns effectively.

In addition to the standard ARIMA parameters, namely *p, d, q*, SARIMA introduces three more parameters, denoted as (*P, D, Q*). These supplementary parameters correspond to the Autoregressive component (P), the Differencing component (D), and the Moving Average Component (Q) at the seasonal level.

The SARIMA model can be formally represented as follows:

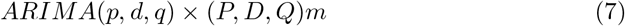

In the context of the SARIMA model, *P* represents the number of seasonal autoregressive terms, *D* signifies the number of seasonal differencing operations, *Q* denotes the number of seasonal moving average terms, and *m* stands for the number of observations in each seasonal cycle. Much like the ARIMA model, we employ the Autocorrelation Function (ACF) and Partial Autocorrelation Function (PACF) to determine the optimal values for these parameters. To evaluate the performance of the developed SARIMA models, we utilize metrics such as Root Mean Square Error (RMSE) and Mean Square Error (MSE).

#### Recurrent Neural Network (RNN)

The Recurrent Neural Network (RNN), a subset of artificial neural networks, was first introduced through David Rumelhart’s work in 1986 [21]. RNNs are equipped with a memory element in the form of a hidden state vector, enabling them to consider information from previous time steps when processing new input. This memory capacity makes RNNs valuable in predicting future values in a time series or detecting underlying patterns. In the domain of time series forecasting, RNNs are employed to make predictions about forthcoming data points by training on historical data. The primary advantage of RNNs in this context lies in their ability to capture temporal dependencies among data points, which often proves crucial in time series forecasting tasks. Recurrent Neural Networks (RNNs) are specialized neural networks designed for processing sequential data, and they have found significant applications in the realm of time series analysis. The fundamental concept behind recurrent neural networks involves incorporating not only the current input data but also leveraging information from previous outputs to make predictions for the present moment. This approach intuitively allows the network to propagate values forward in time. However, implementing such straightforward solutions often proves challenging, as they tend to be difficult to train and exhibit a tendency to forget crucial information. Consequently, a more sophisticated system with a form of memory is essential [19].

Two widely adopted and effective models within the realm of RNNs are Long Short-Term Memory (LSTM) and Gated Recurrent Unit (GRU). These models have demonstrated exceptional performance, overcoming the limitations associated with simpler architectures. In this paper, we utilize Long Short-Term Memory (LSTM).

#### Long Short-Term Memory (LSTM)

Long Short-Term Memory (LSTM), introduced by Hochreiter and Schmidhuber in 1997 [22], represents a gated memory unit within neural networks. Characterized by three gates, LSTM effectively governs the contents of its memory. These gates operate as straightforward logistic functions of weighted sums, with the weights subject to learning through the backpropagation process. Despite its apparent complexity, LSTM seamlessly integrates into neural networks and aligns with their training procedures. It possesses the capability to learn, retain, and recall information without necessitating specialized training or optimization.

The input gate, equation8, and the forget gate, equation9, play pivotal roles in managing the cell state, equation 11, which constitutes the long-term memory component. Simultaneously, the output gate, equation 10 generates the output vector or hidden state, equation 12, representing the memory focused on for utilization. This intricate memory system endows the network with the ability to retain information for an extended duration, a feature notably absent in conventional vanilla recurrent neural networks.

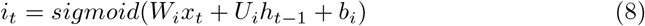

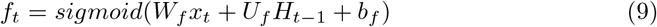

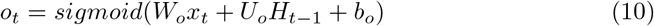

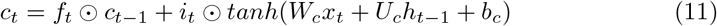

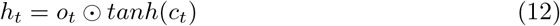

## 3 Results and discussion

In this section, we initially outline the modeling outcomes for confirmed and deceased COVID-19 cases in four states: Alabama, Massachusetts, California, and Washington. We employ the SMA, EWMA, and HWDESA models for this modeling phase. Subsequently, we present the forecasting results for these states using the ARIMA, SARIMA, and RNN models. It is worth noting that the reported case figures may not precisely correspond to the number of individuals who tested positive. Variations can arise due to factors such as untested infected individuals or those who do not seek medical care at healthcare facilities.

### 3.1 Prediction

#### SMA

In our modeling approach, we apply the Simple Moving Average (SMA) to four states using two different parameter settings: 10 days and 30 days. As depicted in Figure 1, a consistent trend emerges across all four states with respect to confirmed cases. Specifically, as the moving average parameter increases, the model’s performance tends to deteriorate. This outcome aligns with our expectations, given the inherent characteristics of the simple moving average method. Furthermore, it is notable that for short-term modeling, the Simple Moving Average proves to be sufficiently effective.

**Fig. 1.**
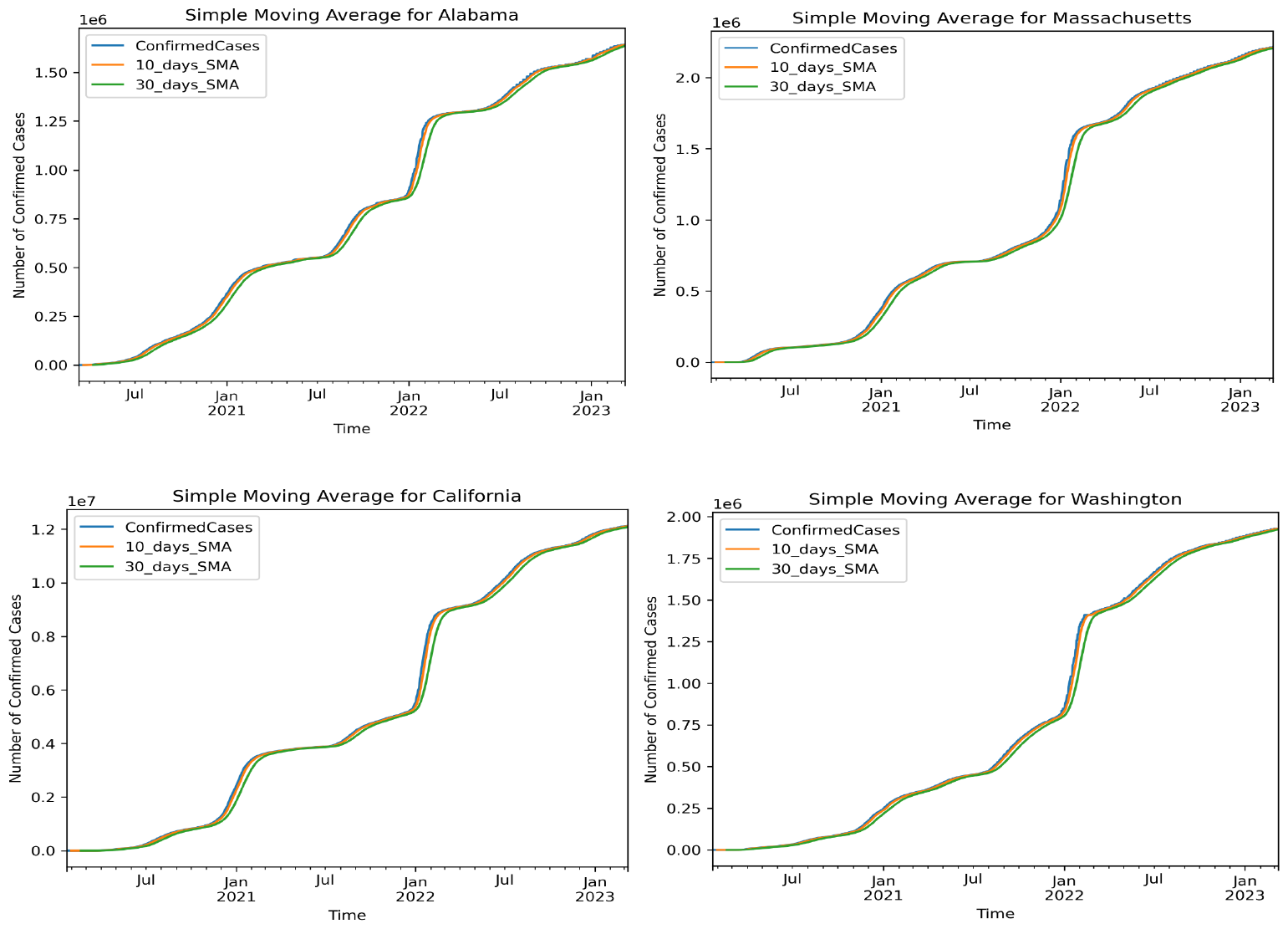
Modeling the number of confirmed cases with SMA model in four states

#### EWMA

For the calculation of the Exponentially Weighted Moving Average (EWMA), we utilize 30 days of recent data. The results of the EWMA method are presented in Figure 2 using a 30-day parameter setting.

**Fig. 2.**
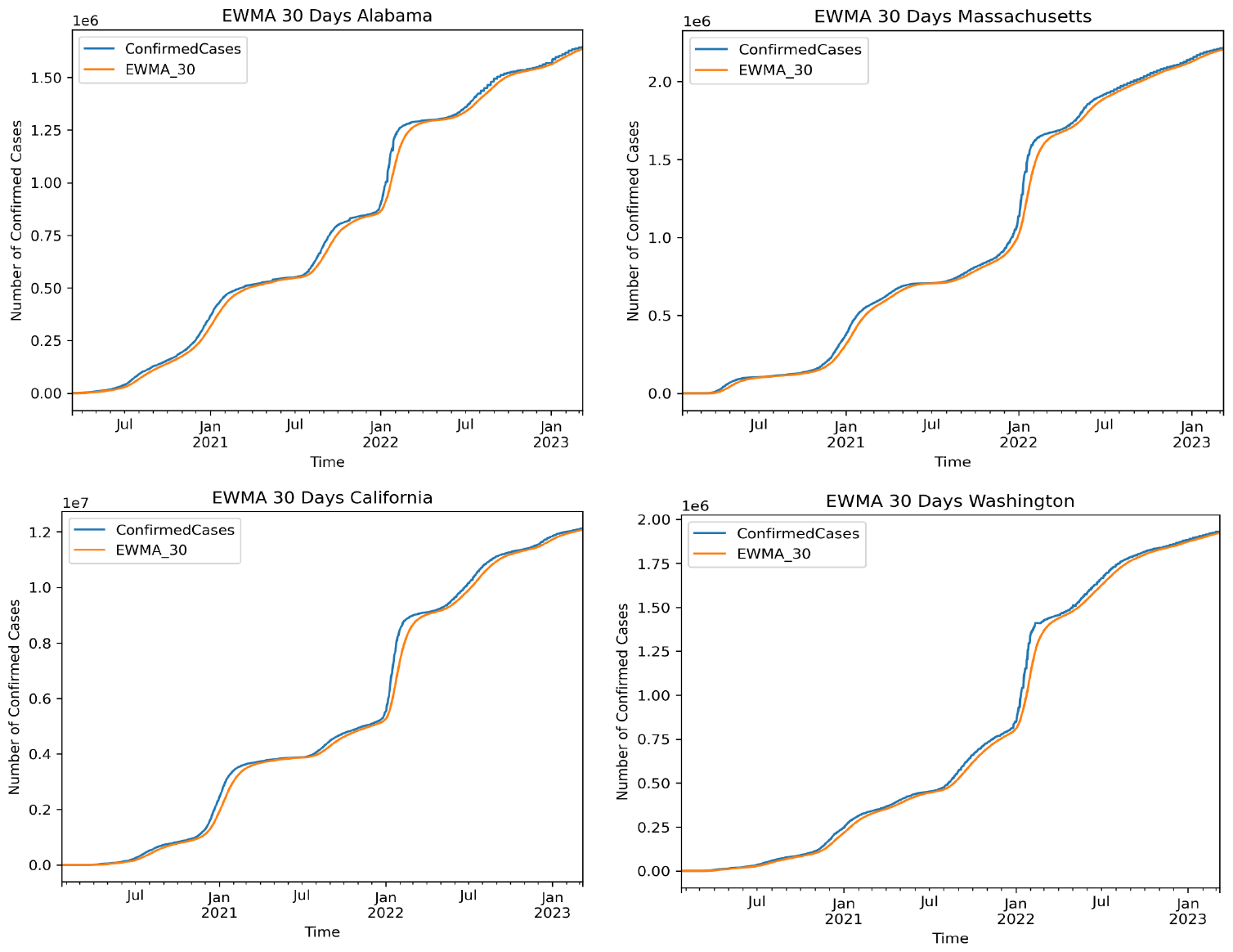
Modeling the number of confirmed cases with EWMA model in four states

Notably, akin to the SMA approach, the EWMA method consistently underestimates the number of confirmed cases in all four states. This pattern indicates a degree of bias in the methodology for these specific data points.

#### Holt-Winters Double Exponential Smoothing Additive

Our analysis underscores the effectiveness of the Holt-Winters Double Exponential Smoothing model as a robust representative among models designed to handle both trend and seasonality in time-series data. It’s worth noting that although the seasonal variations within the data appear relatively consistent throughout the series, we still explore both additive and multiplicative models.

Upon evaluating model performance using the Root Mean Square Error (RMSE) metric, we draw the conclusion that the additive model surpasses the multiplicative model when applied to COVID-19 data. In both models, we set the span to 30 days, and the smoothing parameter *α* is computed as 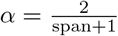. Figure 3 provides a visual representation of the performance comparison, illustrating that the Holt-Winters (HW) model outperforms the Exponentially Weighted Moving Average (EWMA) with a 30-day parameter setting. Specifically, the EWMA consistently underestimates the number of confirmed cases, whereas the HW model almost impeccably captures the case counts.

**Fig. 3.**
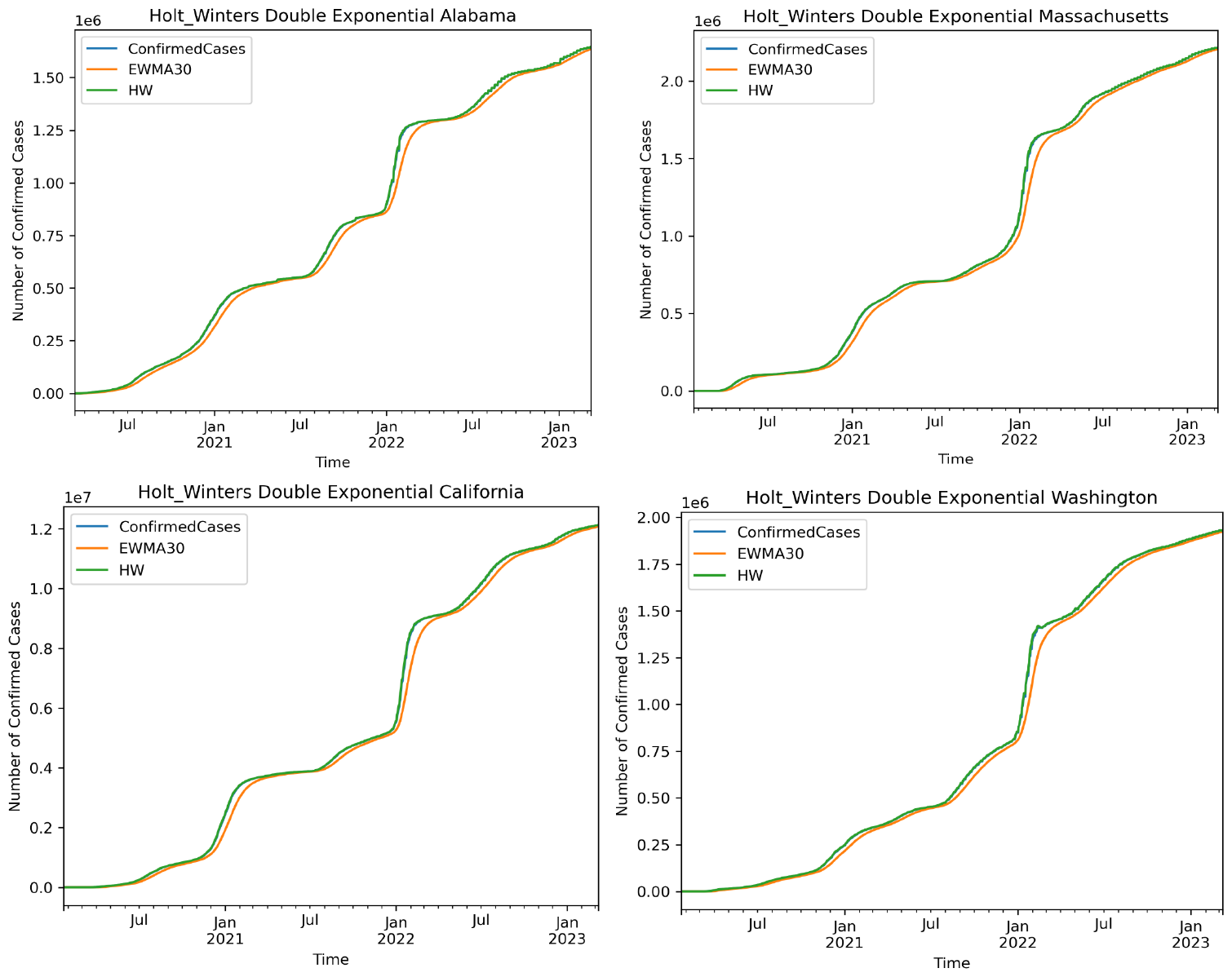
Modeling the number of cases with Holt-Winters Double Exponential Smoothing in four states

### 3.2 Forecasting

#### ARIMA Model

For the forecasting phase, we extend our predictions for a duration of 90 days. By comparing the Root Mean Square Error (RMSE), as illustrated in Figure 4, we find that the ARIMA model outperformed in predicting the number of confirmed cases in Alabama, California, Massachusetts, and Washington, respectively.

**Fig. 4.**
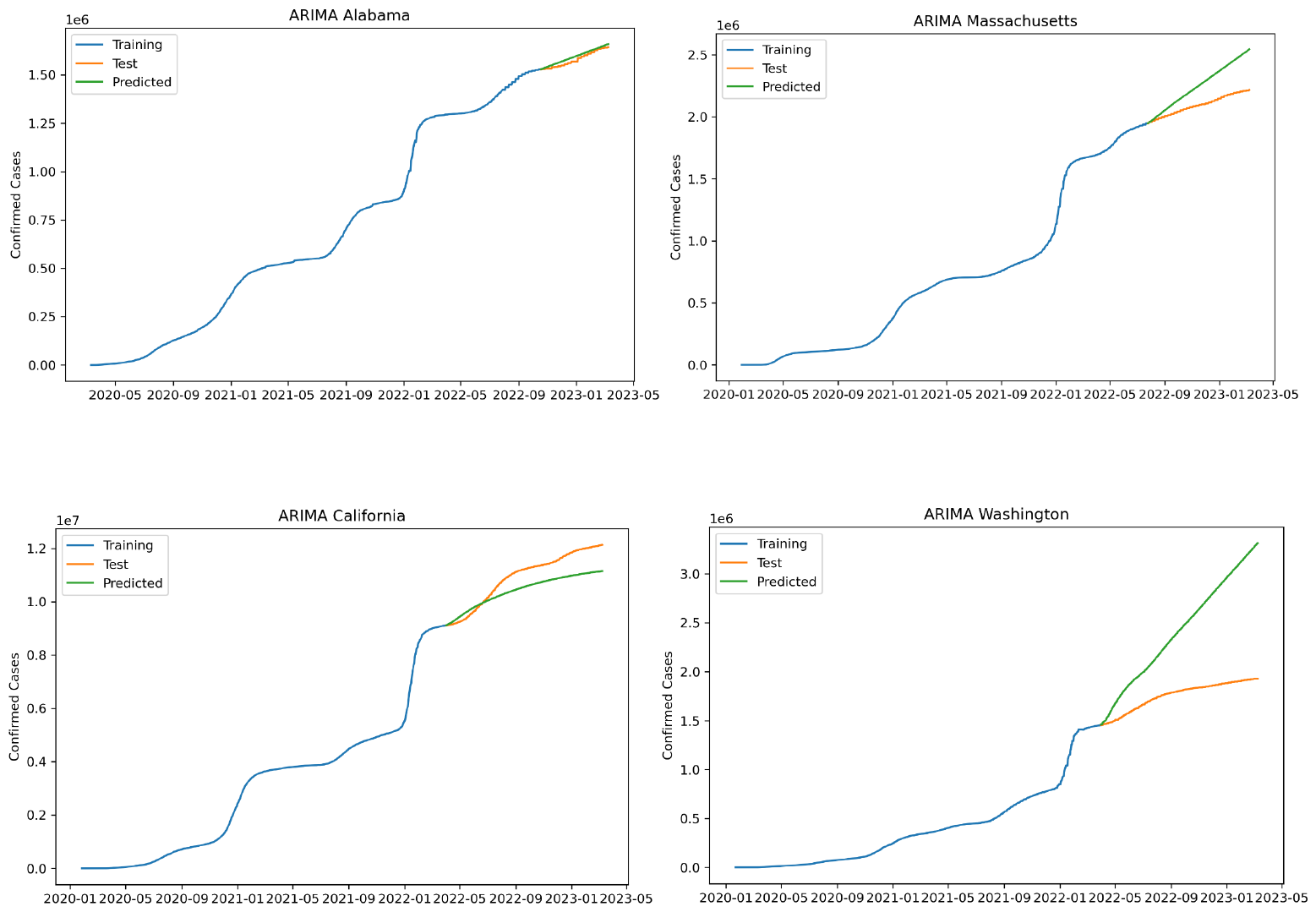
Forecasting the number of cases with ARIMA model in four states

It is important to note that the model’s performance varies across different states. To optimize the ARIMA model, we adopt an iterative approach to explore various combinations of parameters (p, d, and q). Allow us to elucidate this procedure: the grid search systematically identifies the best combination that minimizes the Akaike Information Criterion (AIC). In each iteration, the ARIMA model is generated for the training dataset, and the grid search stores the lowest error for each iteration. The iterative process continues until ten successive models fail to outperform the lowest error previously achieved. This way, we ensure that the parameters selected are robust and reliable for forecasting.

Table 1 represents the ARIMA parameters and RMSE for each state.

**Table 1.** The ARIMA model’s parameters and RMSE for each state.

| State | Parameters | RMSE |
| --- | --- | --- |
| Alabama | ARIMA (0,1,1) | 16437.64 |
| Massachusetts | ARIMA (2,1,0) | 190768.30 |
| California | ARIMA (7,1,2) | 628903.03 |
| Washington | ARIMA (0,1,0) | 766150.46 |

#### SARIMA Model

As with the ARIMA model, we extend our forecasting horizon to 90 days using the Seasonal Autoregressive Integrated Moving Average (SARIMA) model. The performance of this model for each state is vividly portrayed in Figure 5. After meticulous evaluation of the Root Mean Square Error (RMSE), the SARIMA model stands out as a superior predictor for the number of cases in all four states.

**Fig. 5.**
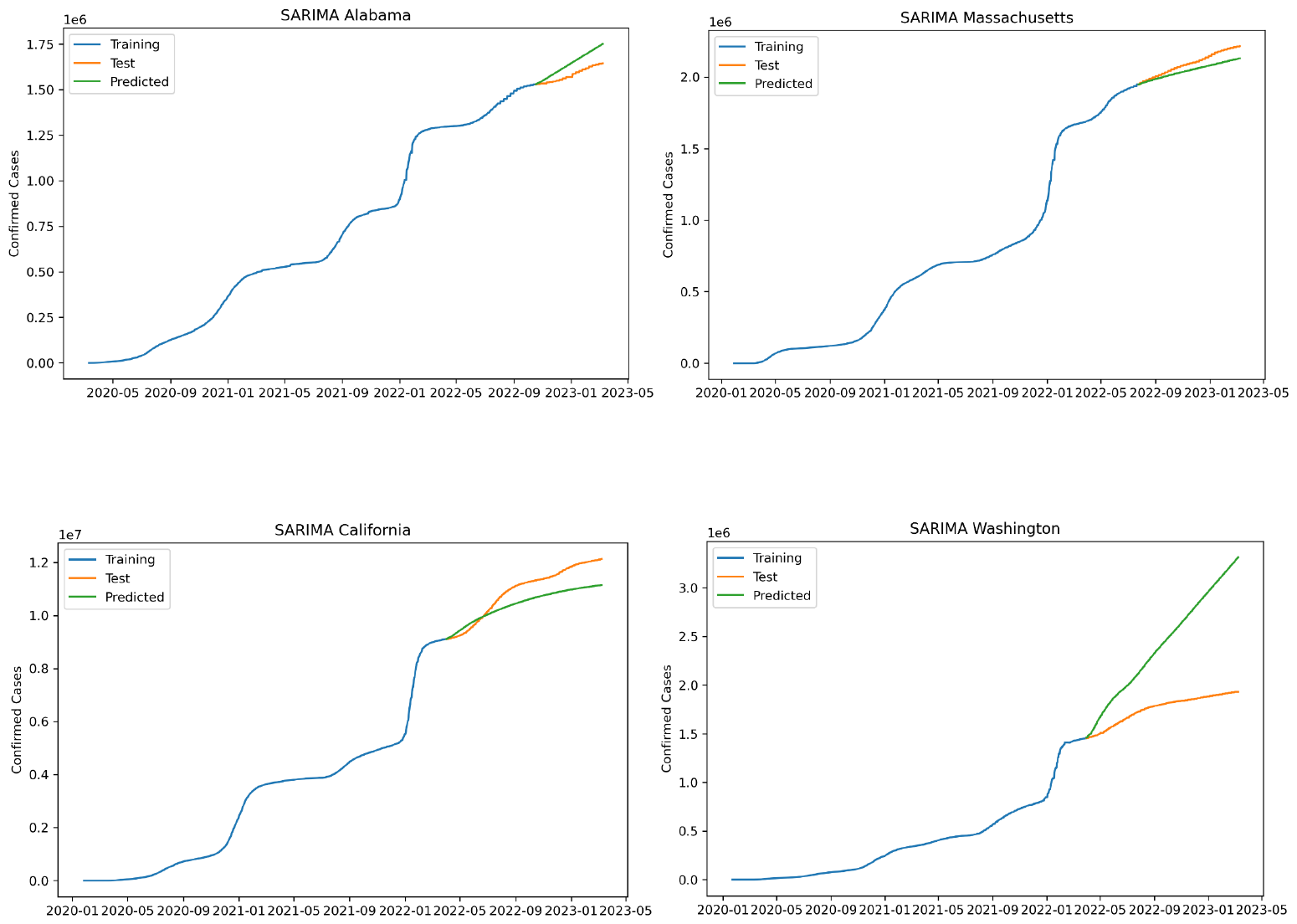
Forecasting the number of cases with SARIMA model in for states

Table 2 illustrates the SARIMA parameters and RMSE for each state. Table 2 illustrates the SARIMA parameters and RMSE for each state.

**Table 2.**
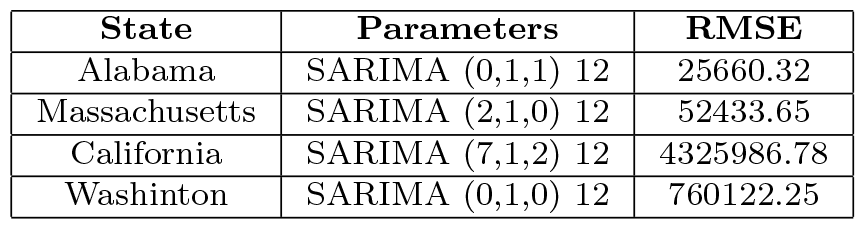
The SARIMA model’s parameters and RMSE for each state.

| State | Parameters | RMSE |
| --- | --- | --- |
| Alabama | SARIMA (0,1,1) 12 | 25660.32 |
| Massachusetts | SARIMA (2,1,0) 12 | 52433.65 |
| California | SARIMA (7,1,2) 12 | 4325986.78 |
| Washington | SARIMA (0,1,0) 12 | 760122.25 |

#### RNN

Figure 6 provides a comprehensive representation of the performance of the Recurrent Neural Network (RNN) methodology in predicting the number of cases across the same four states. The results notably demonstrate the substantial superiority of the RNN model in forecasting the number of confirmed cases within the states under examination, outperforming both the ARIMA and SARIMA models by a significant margin.

**Fig. 6.**
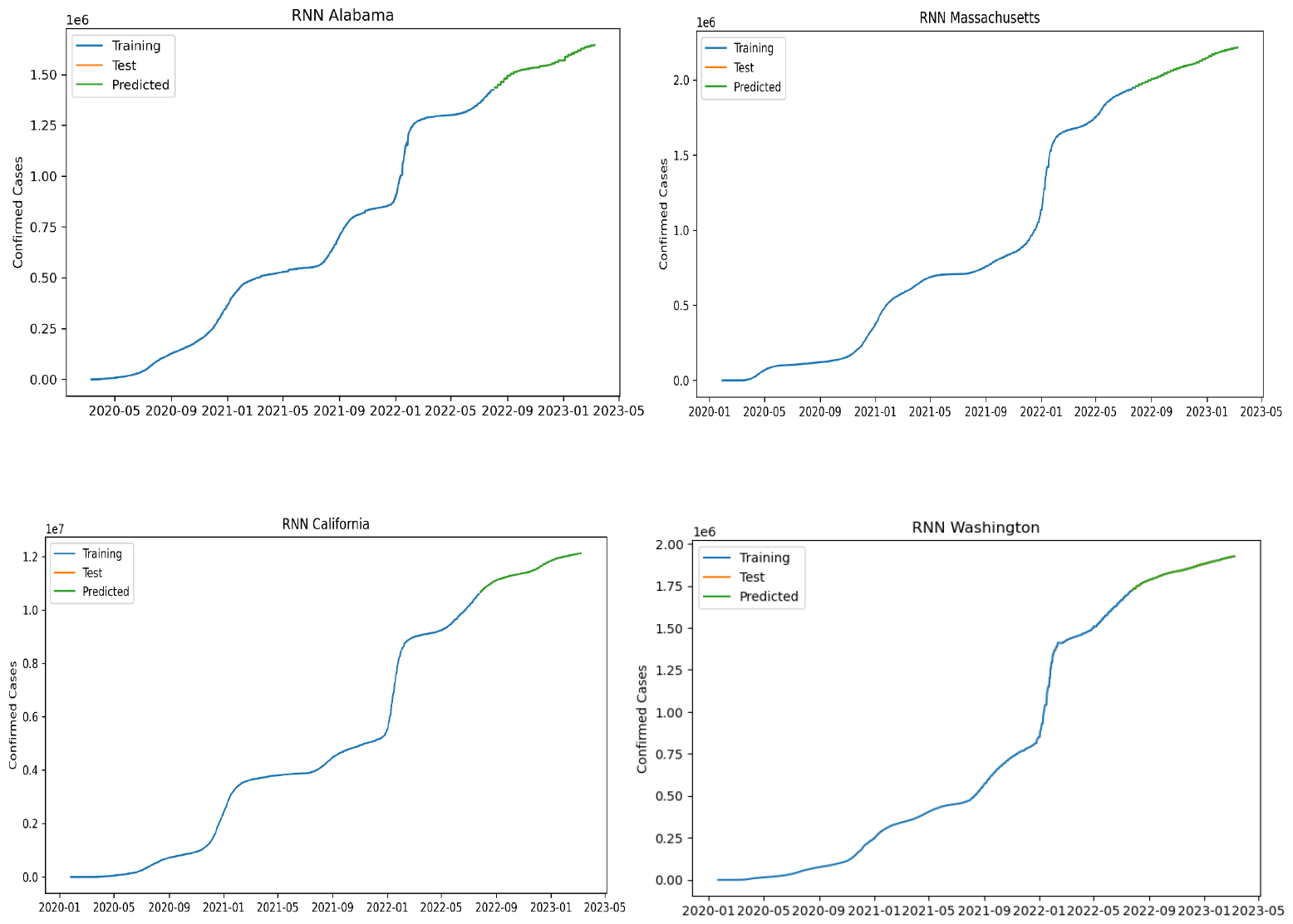
Forecasting the number of cases with RNN in four states

## Conclusion

Time-series techniques have played a significant role in modeling data associated with temporal patterns. In this paper, we have applied three fundamental models, namely Simple Moving Average, Exponentially Weighted Moving Average, and Holt-Winters

Double Exponential Smoothing Additive, to model the counts of confirmed COVID-19 cases. Additionally, we have harnessed the power of advanced techniques such as ARIMA, SARIMA, and RNN for both modeling and forecasting the number of cases. These models serve as valuable tools for governors and healthcare providers, facilitating effective management, planning, and preparedness for the peaks of COVID-19 cases and similar infectious diseases.

In our evaluation, we have leveraged RMSE and MSE measures to compare the performance of these methods in modeling and forecasting case counts. Notably, the results indicate that among the three fundamental models, the Holt-Winters Double Exponential Smoothing Additive model consistently outperforms the other two in estimating the number of infected individuals. Similarly, the RMSE measure demonstrates that the RNN model excels as a superior forecaster of infected cases in comparison to both ARIMA and SARIMA.

However, it’s essential to acknowledge certain research limitations. One limitation pertains to the relatively limited number of independent variables in the dataset, where only the time series of case counts is considered. There exists potential for other explanatory variables (EV) to influence the number of COVID-19 cases, such as individuals’ medical history or preexisting health conditions. Incorporating such variables into the modeling framework could enhance forecasting accuracy. Should such data become available, models like ARIMAX and SARIMAX, designed to incorporate additional explanatory variables in time series models, can be employed. Furthermore, the exploration of advanced machine learning methods, including deep learning techniques, holds the potential to further enhance forecasting accuracy and capture intricate patterns within the data.

## Data Availability

The Johns Hopkins COVID-19 data repository is widely acknowledged as the most
reliable source of information for COVID-19 statistics

https://coronavirus.jhu.edu/map.html

## Notes

### Competing Interest Statement

The authors have declared no competing interest.

### Funding Statement

No funding was received for this research

